# Elevated HScore is Associated with Poor Clinical Outcomes in COVID-19, Even in the Absence of Secondary Hemophagocytic Lymphohistiocytosis

**DOI:** 10.1101/2021.01.26.21249335

**Authors:** Rafael Benavente, Camila Peña, Allyson Cid, Nicolás Cabello, Pablo Bustamante, Marco Álvarez, Elizabeth Henríquez, Andrés Soto, Erika Rubilar

## Abstract

**Introduction:** Patients with Coronavirus Disease 2019 (COVID-19) frequently experience a hyperinflammatory syndrome, that leads to unfavorable outcomes. This condition resembles Secondary Hemophagocytic Lymphohistiocytosis (sHLH) described in neoplastic, rheumatic and other infectious diseases. However, it has not been prospectively studied on these patients. A scoring system (HScore) has been validated for sHLH, and recently proposed to evaluate hyperinflammation in COVID-19.

**Methods:** 143 patients aged ≥18 years admitted because of COVID-19 were enrolled in a prospective, single-center, cohort study. HScore was calculated within the 72 hours since admission. The incidence of sHLH during hospitalization was evaluated. Additionally, the relationship between HScore ≥130 points and either the requirement of mechanical ventilation or 60-days mortality was explored.

**Results:** The median age of enrolled patients was 57 (21-100), and 63.6% were male. The median HScore was 96 (33-169). One patient was diagnosed with sHLH (incidence 0,7%), due to a HScore of 169. After adjusting for age, sex, comorbidities and obesity, HScore ≥130 was independently associated with the composite clinical outcome (HR 2.13, p=0.022).

**Conclusion:** sHLH is not frequent among COVID-19 patients. HScore can efficiently predict the risk for poor outcomes.

## Introduction

Since the beginning of the severe acute respiratory syndrome coronavirus 2 (SARS-CoV-2) pandemic, it has been noticed that patients frequently undergo a hyperinflammatory syndrome, contributing to worse outcomes [1]. Addressing this issue is important due to the potential benefit of immunomodulatory therapies. Many aspects of this syndrome resemble Secondary Hemophagocytic Lymphohistiocytosis (sHLH) that has been described in malignancies, rheumatologic conditions and infectious diseases, including several caused by viruses [2-3]. sHLH is characterized by high fever, cytopenias, organomegaly, elevated inflammatory markers, and frequently (though not always) by hemophagocytosis in bone marrow or lymph nodes [4]. Once presented, sHLH is associated with high mortality and aggressive therapy must be initiated in order to control the hyperinflammatory state. Treatments are usually complex and evidence-based data regarding the best approach remain scarce [5-6].

The HLH-2004 study, established the first commonly clinical criteria used for diagnosis of sHLH [7]. More recently, a clinical scoring system (HScore) has been developed and validated for the diagnosis of sHLH [8], with an optimal cutoff value of 169 points to predict the disease.

HScore has also been proposed for measuring hyperinflammation in COVID-19 patients [9]. Thus, the aim of this study was to assess the presence of sHLH among patients with COVID-19 admitted for hospitalization in one center in Santiago (Chile), and to evaluate HScore as a prognostic tool for poor outcomes.

## Patients and Methods

### Study design

Patients aged 18 years and older hospitalized because of COVID-19, between April 1st and May 31st of 2020, were considered for enrollment into a prospective, single-center, cohort study at the Hospital del Salvador in Santiago, Chile.

SARS-CoV-2 infection was confirmed using real-time polymerase chain reaction (rt-PCR) from nasopharyngeal swabs in all cases. Patients with asymptomatic SARS-CoV-2 infection (eg. pre-surgical testing), or COVID-19 cases in which the infection was suspected to be acquired during hospitalization, were excluded.

The primary objective of this study was to assess the incidence of sHLH among hospitalized COVID-19 patients. A secondary objective, was to evaluate the relationship between elevated HScore and a composite endpoint comprising mechanical ventilation and 60-days mortality from any cause, in a multivariate analysis.

### Data collection

Within 72 hours since admission, HScore was calculated for every enrolled patient, including the following parameters: history of known immunosuppression, highest body temperature registered, number of cytopenias, organomegaly, concentration of ferritin, triglycerides, fibrinogen and aspartate aminotransferase (AST). Immunosuppression was defined as being HIV positive or receiving long-term immunosuppressive therapy (eg. glucocorticoids, cyclosporine, azathioprine). Cytopenia was defined as either haemoglobin concentration <9,2 g/dL, a white blood cell count <5000 cells per mm3, or platelet count <110.000 per mm3. Organomegaly was defined as hepatomegaly and/or splenomegaly based on physical examination or imaging studies. We also recorded routine demographic and clinical data. Comorbidities were assessed by the Charlson Comorbidity index (CCI), as it has been previously reported in this population [10]. Multiple comorbidities were defined as having two or more points in CCI. Obesity was defined as a body mass index (BMI) of 30 Kg/m2 or more.

### sHLH diagnosis and HScore as a prognostic tool

Once HScore was assessed, if sHLH was clinically suspected, haematological consultation was requested for further evaluation. Bone marrow aspirate or biopsy were performed only when clinically indicated, and whether it was considered useful for the patient management. We considered a definitive diagnosis of sHLH when HScore was equal or superior to 169 points, in absence of an alternative diagnosis.

For prognostic purposes, we defined “high” HScore as having 130 points or more as this cut-off has been reported previously, anticipating difficulties to perform a bone marrow analysis in critical care patients or for security reasons [11,12].

### Statistical analysis

Continuous variables are presented as median (range) and categorical variables as n (%). Comparisons between high or low HScore groups were made using Student’s t test, Chi-square (X^2^) or Mann-Whitney *U* test, as appropriate. The incidence of sHLH was calculated as the number of sHLH cases to the total enrolled patients. Mortality from any cause was evaluated until 60 days since admission. For the multivariate analysis, a Cox proportional hazards model was used including other reported risk factors for worse outcomes: age, male sex, multiple comorbidities, and obesity.

The study protocol received ethical approval from the institutional ethics committee (Comité de Ética Científica del Servicio de Salud Metropolitano Oriente. Santiago, Chile).

## Results

### Population Characteristics and sHLH frequency

From april 1^st^ to may 31^sth^ 2020, 143 patients were evaluated. The median age was 57 years (21-100), and 91 (63.6%) were male. The median time of symptoms before admission was 7 days (1-40). The median CCI was 1 (range 0-9) and 43 (30,1%) patients were obese. Seventy-one patients (49,7%) were admitted to Intensive Care Units, and 42 (29,4%) needed mechanical ventilation. Twenty-two (15,4%) patients received corticosteroids. Thirty-six (25,2%) patients died during follow-up (Table 1).

**Table 1.** Cohort Demographic and Clinical Characteristics.

|  | Whole Cohort<br>(n=143) | HScore <130<br>(n=121) | HScore ≥130<br>(n=22) | p<br>value |
| --- | --- | --- | --- | --- |
| **Age (median,range)** | 57 (21-100) | 64 (21-100) | 59 (28-78) | 0.105 |
| **Male sex (n, %)** | 91 (63,6%) | 70 (57,9%) | 21 (95,4%) | 0.001* |
| **Charlson Comorbidity Index (median,range)** | 1 (0-9) | 1 (0-7) | 0 (0-9) | 0.236 |
| *Acute myocardial infarction (n, %) | 9 (6,3%) | 3 (2,5%) | 6 (27,2%) |  |
| *Congestive heart failure (n, %) | 7 (4,9%) | 7 (5,8%) | 0 (0%) |  |
| *Peripheral vascular disease (n, %) | 2 (1,4%) | 2 (1,7%) | 0 (0%) |  |
| *Cerebrovascular disease (n, %) | 1 (0,7) | 1 (0,8%) | 0 (0%) |  |
| *Hemiplegia (n, %) | 0 (0%) | 0 (0%) | 0 (0%) |  |
| *Dementia (n, %) | 13 (9,1%) | 13 (10,7%) | 0 (0%) |  |
| *Chronic lung disease (n, %) | 25 (17,5%) | 22 (1,8%) | 3 (13,6%) |  |
| *Rheumatic disease (n, %) | 7 (4,9%) | 6 (4,9%) | 1 (4,5%) |  |
| *Peptic ulcer (n, %) | 0 (0%) | 0 (0%) | 0 (0%) |  |
| *Mild liver disease (n, %) | 0 (0%) | 0 (0%) | 0 (0%) |  |
| *Severe liver disease (n, %) | 2 (1,4%) | 2 (1,7%) | 0 (0%) |  |
| *Diabetes without end organ damage (n, %) | 29 (20,3%) | 27 (22,3%) | 2 (9,1%) |  |
| *Diabetes with End organ damage (n, %) | 12 (8,4%) | 10 (8,3%) | 2 (9,1%) |  |
| *Moderate to Severe kidney disease (n, %) | 9 (6,3%) | 7 (0,6%) | 2 (9,1%) |  |
| *Solid malignancy, nonmetastatic (n, %) | 8 (5,6%) | 7 (5,8%) | 1 (4,5%) |  |
| *Solid malignancy, metastatic (n, %) | 1 (0,7%) | 0 (0%) | 1 (4,5%) |  |
| *Leukemia (n, %) | 1 (0,7%) | 1 (0,8%) | 0 (0%) |  |
| *Lymphoma (n, %) | 1 (0,7%) | 1 (0,8%) | 0 (0%) |  |
| *AIDS (n, %) | 0 (0%) | 0 (0%) | 0 (0%) |  |
| **Body Mass Index ≥ 30 Kg/m2 (n, %)** | 43 (30,1%) | 37 (30,6%) | 6 (27,3%) | 0.685 |
| **Days of symptoms at admission (median, range)** | 7 (1-40) | 7 (1-40) | 8 (1-15) | 0.740 |
| **Corticosteroid treatment (n %)** | 22 (15,4%) | 15 (12,4%) | 7 (31,8%) | 0.020* |
| **Critical Care admission (n, %)** | 71 (49,7%) | 54 (44,6%) | 17 (77,2%) | 0.005* |
| **Mechanical Ventilation (n, %)** | 42 (29,4%) | 28 (23,1%) | 14 (63,6%) | 0.000* |
| *Mechanical ventilation days (median, range) | 11 (1-37) | 11(1-37) | 11 (5-23) | 0.902 |
| **Hospital Length of Stay in Days (median, range)** | 9 (1-72) | 8 (1-62) | 16 (1-72) | 0.002* |
| **60-days Mortality** | 36 (25,2%) | 27 (22,3%) | 9 (40,9%) | 0.065 |

The median HScore was 96 (33-169). One patient was diagnosed with sHLH, due to a HScore of 169 points (incidence 0,7%). He was a male with a high-grade astrocytoma. Due to poor prognosis, no further immunosuppressive therapy was given, passing five days after admission. After initial scoring, sHLH was clinically suspected in four additional patients. Three of these patients had advanced cancer, and one had a liver transplant. In one of them, the diagnosis was ruled out after bone marrow biopsy; whereas the remaining patients were not biopsied because it was not considered useful for patient management (terminally ill).

### HScore as prognostic factor

After dividing the patients in low versus high HScore groups, we observed some significant differences, with the latter group being more likely to be male, admitted to ICU or prescribed corticosteroids.

Fifty-eight patients experienced the composite endpoint during follow-up. In the multivariate analysis, and after adjusting for age, sex, comorbidities and obesity, Hscore ≥130 points was independently associated with the composite endpoint (HR 2.13, CI 1.18 – 4.06, p 0.022) (Table 2).

**Table 2.** Multivariate analysis (n=143)

|  | HR | CI | p value |
| --- | --- | --- | --- |
| Age (years) | 1,03 | 1,015 - 1,055 | <0.001* |
| Male sex | 1,42 | 0,766 - 2,638 | 0,264 |
| Multiple Comorbidities (CCI≥2) | 1,67 | 0,943 - 2,943 | 0,079 |
| Obesity (BMI ≥ 30 Kg/m<sup>2</sup>) | 2,19 | 1,256 - 3,807 | 0,006* |
| HScore ≥130 | 2,13 | 1,117 - 4,061 | 0,022* |

CCI: Charlson Comorbidity Index. BMI: Body Mass Index

## Discussion

To the best of our knowledge, this is the first report that prospectively followed eventual sHLH in hospitalized patients with COVID-19. sHLH was found to be uncommon in these patients, suggesting other causes of hyperinflammation.

### sHLH in COVID-19

Several groups have reported high ferritin in patients with severe COVID-19 [13-14]. This finding, associated with other markers of hyperinflammation, led to the proposal of the association with sHLH [7,15]. In previous viral outbreaks, the role of sHLH was also suggested. In the SARS-associated coronavirus (SARS-CoV) epidemic, hemophagocytosis were reported in some autopsies [16]. In the Middle East respiratory syndrome coronavirus (MERS□CoV) outbreak, a sHLH-like syndrome was also noticed [17].

The pathophysiology of sHLH is not fully understood. It is frequently associated with malignancies, infections or inflammatory/rheumatic diseases. In some patients, these conditions can trigger a cytotoxic T lymphocytes-driven macrophage activation, leading to hypercytokinemia, which clinically presents as fever, cytopenias and organ failure. Virus-associated sHLH occurs commonly among immunosuppressed patients but also in healthy hosts. The most common virus infection triggering sHLH is Epstein-Barr Virus (EBV) [2]. However, sHLH is not frequently associated with respiratory failure, as seen among SARS-CoV2 patients. Rather, sHLH produces organomegalies such as hepatosplenomegaly or lymphadenopathy.

Our results suggest that sHLH is not behind the hyperinflammation state of COVID-19. Within the whole cohort, only one patient fulfilled sHLH criteria during early hospitalization; whereas in only four additional patients sHLH was suspected and excluded thereafter during evolution. Interestingly, all those patients had predisposing pathologies; high-grade astrocytoma, advanced breast cancer (one patient), lymphoma (two patients), and recent liver transplantation (one patient). This observation raises the question whether these patients had indeed sHLH manifestations due to COVID-19, or related to comorbidities largely known to be associated with sHLH.

Cytopenias occurs in 60-70% of patients with sHLH [2]. However, very few COVID-19 patients presented cytopenias, and if they did, HLH-2004 or HScore criteria were not met. Most of the patients also presented high ferritin levels, which reflects inflammation (after excluding multiple red blood cell transfusions). Fibrinogen was elevated in most COVID-19 patients, contrary to what we should expect in sHLH. This may be reflecting the hypercoagulability state of this disease, and the fact that fibrinogen is an acute-phase protein. It would have been desirable to measure IL-6 levels, since it is modestly elevated in sHLH unlike other hyperinflammatory syndromes, as Cytokine Release Syndrome (CRS) described in Chimeric antigen receptor T (CAR-T) cells or Haploidentical Hematopoietic Stem Cell Transplantation (HSCT) [6]. Furthermore, there have been reports of high levels of IL-6 in COVID-19, but again, not as high as seen in CRS [18]. This observation has led some investigators to propose hyperinflammation in this pathology does not correspond to neither CRS nor sHLH [19], and might be similar to the so-called Macrophage Activation-Like Syndrome in sepsis [20-21].

### HScore as a predictor of worse outcomes

Inflammation is a major cause of morbility and mortality among COVID-19 patients, as previously reported in infections by other coronaviruses [22]. This can be explained by an uncontrolled, self-perpetuating, and tissue-damaging inflammatory activity, highlighted by the increase of a number of markers of inflammation in the curse of the disease [23-24]. The observation that clinical worsening is often seen at day 8-10 since the symptoms onset, reinforces the idea that it is hyperinflammation and not the virus itself that produces the most severe manifestations [11]. Several prognostic factors are described so far for this new disease, including age, diabetes, hypertension, cardiovascular disease, obesity, high CCI, D-Dimer and ferritin levels [10, 25-28].

HScore was developed in 2014, to predict the presence of sHLH [6]. Nevertheless, since high HScore at time of hospitalization may reflect severe inflammation rather than sHLH itself, some authors have suggested to use this score in all patients with COVID-19 [8,15]. Although others discourage this approach [29-30], our results suggest 130 or more points of HScore are associated with worse outcomes, namely, the requirement of mechanical ventilation or/and death, in hospitalized patients with COVID-19. HScore is relatively easy to calculate, and can eventually lead to aggressive therapies, such as, more potent immunosuppressive agents or immunomodulators [15,31-34].

There are some limitations in our study. First, we did not have access to all tests listed in sHLH diagnostic criteria. Namely, NK-lymphocytes study is not currently available in our country; while the soluble CD25 (sCD25) study is not available in our institution. As previously stated, bone marrow study was not performed in all patients, to minimize the researchers’ exposure to the virus, and because it was not considered useful for patient management in most cases. Nevertheless, we believe that we can reasonably rule out sHLH as a cause of hyperinflammation in our patients. Finally, during the data collection, hydroxychloroquine or lopinavir/ritonavir were commonly prescribed, while steroids were not. Data from large RCT have modified this practice [35-36] in favour of steroid use, which in turn could affect the power of HScore to predict poor outcomes.

### Conclusion

In COVID-19 patients, sHLH seems to be a rare event, but excessive inflammation is common. In our cohort, high HScore (≥130), even at a lower threshold than required for sHLH diagnosis, was associated with poor outcomes. Further studies validating this finding could be helpful to select patients for more aggressive immunosuppressive treatments.

The authors declare that they have no conflict of interest.

## Data Availability

All data referred to in the manuscript are available.

